# The safety of antivirals and neutralising monoclonal antibodies used in prehospital treatment of Covid-19

**DOI:** 10.1101/2024.02.19.24303044

**Authors:** Katie Bechman, Amelia Green, Mark D Russell, Zijing Yang, Bang Zheng, Sam Norton, Rebecca M Smith, Amir Mehrkar, Sebastian Bacon, Ben Goldacre, Brian MacKenna, James Galloway, OpenSAFELY Collaborative

## Abstract

**Objective:** This proof-of-principle pharmacovigilance study used Electronic Health Record (EHR) data to examine the safety of sotrovimab, paxlovid and molnupiravir in prehospital treatment of Covid-19.

**Method:** With NHS England approval, we conducted an observational cohort study using OpenSAFELY-TPP, a secure software-platform which executes analyses across EHRs for 24 million people in England. High-risk individuals with Covid-19 eligible for prehospital treatment were included. Adverse events (AEs) were categorised into events in the drug’s Summary of Product Characteristics (SmPC), drug-reactions and immune-mediated. Cox models compared risk across treatments. A pre-pandemic record analysis was performed for comparative purposes.

**Results:** Between 2021-2023, 37,449 patients received sotrovimab, paxlovid or molnupiravir whilst 109,647 patients made up an eligible-but-untreated population. The 29-day rates of AEs were low: SmPC 0.34 per 1000 patient-years (95%CI 0.32-0.36); drug-reactions 0.01(95% CI0.01-0.02) and immune-mediated 0.03(95%CI 0.03-0.04), and similar or lower than the pre-pandemic period. Compared with the eligible but untreated population, sotrovimab and paxlovid associated with a risk of SmPC AE [adjHR 1.36(95%CI 1.15-1.62) and 1.28(95%CI 1.05-1.55), respectively], whilst sotrovimab associated with a risk of drug-reactions [adjHR 2.95(95%CI 1.56-5.55)] and immune-mediated events [adjHR 3.22(95%CI 1.86-5.57)].

**Conclusion:** Sotrovimab, paxlovid and molnupiravir demonstrate acceptable safety profiles. Although the risk of AEs was greatest with sotrovimab, event rates were lower than comparative pre-pandemic period.

## Introduction

The use of antivirals and neutralising monoclonal antibodies (nMAB) in the prehospital treatment of Covid-19 has the potential to improve outcomes for high-risk individuals. This has been demonstrated in early randomised control trials (1-3) and observational studies (4-6), although not supported by a more recent open label trial in a highly vaccinated population (7).

In the UK, intravenous nMAB, sotrovimab, and oral antivirals, paxlovid (nirmatrelvir/ritonavir) and molnupiravir, have been approved for use. The Medicines and Healthcare products Regulatory Agency (MHRA) granted conditional marketing authorisation for emergency use of these therapies, as the benefit of immediate drug availability outweighed the risks of a less vigorous standard approval. Since December 2021, Covid-19 medicine delivery units (CMDUs) have been providing treatment with sotrovimab, paxlovid and molnupiravir for individuals at high risk of developing severe Covid-19 outcomes in community settings across England (8, 9). Lower treatment coverage has been observed in certain groups, in particular: different NHS regions, minority ethnic groups, people aged ≥80 years, those living in socioeconomically deprived areas, and care home residents. (10)

The safety data from the clinical trial programme for all three drugs has been largely reassuring (1-3). However, these randomised control trials are limited in their ability to robustly examine adverse events (AEs): they are powered to determine drug efficacy with a far smaller sample size than that required to detect rarer side effects; they are often short in duration, potentially missing events that have a lagged exposure risk; and they self-select for patients who are fit enough to participate in research, often recruiting a healthier cohort that is not representative of the general population. Also of relevance is the definition of serious adverse events (SAEs), encompassing any AE that results in inpatient hospitalisation. As the key objective of Covid-19 prehospital treatment is the prevention of admission with Covid-19, it is possible that a reduction in hospitalisation as a result of drug efficacy may bias the safety profile in favour of drug over placebo. This may have impacted safety analyses that report AEs by ‘any cause for hospitalisation’.

The objective of this study was to use real-world data to detect AEs that may have been missed in clinical trials due to homogeneous samples and limited person-years of follow up. This was a proof of principle study using near real-time electronic health record data in the OpenSAFELY-TPP database to examine pharmacovigilance of antivirals and nMABs used prehospital in the prevention of severe Covid-19 outcomes.

## Patients and methods

### Study design

We conducted a retrospective cohort study between December 11^th^ 2021 (the earliest date a patient could have tested positive for SARS-CoV-2 and be eligible for treatment) until June 26^th^ 2023 (the end date for access to community treatments via CMDUs).

### Data source

This analysis was conducted using the OpenSAFELY-TPP database. OpenSAFELY is a highly secure, transparent, open-source software platform that was created on behalf of NHS England for the purpose of urgent Covid-19 research. It executes analyses within the electronic health record vendor’s highly secure data centre. There is access to pseudonymised data across primary care records for patients currently registered with GP surgeries that use TPP SystmOne software, covering 24 million people (40% of the English population). All data were linked, stored, and analysed securely using the OpenSAFELY platform (https://www.opensafely.org/) as part of the NHS England OpenSAFELY Covid-19 service. Data include pseudonymised data, such as coded diagnoses, medications, and physiological parameters. No free text data are included. All code is shared openly for review and re-use under MIT open licence [https://github.com/opensafely/Safety-Sotrovimab-Paxlovid-Molnupiravir]. Detailed pseudonymised patient data is potentially re-identifiable and therefore not shared.

Primary care records managed by the GP software provider, TPP were linked to other pseudonymised datasets, including i) national coronavirus testing records from the Second Generation Surveillance System, ii) accident and emergency and inpatient hospital records from the Secondary Uses Service (SUS) and iii) Covid-19 therapeutics dataset, a patient-level dataset on nMABs and antiviral treatments derived from Blueteq software that CMDUs use to notify NHS England of Covid-19 treatments

### Study population

We included all individuals aged 18 years or older and who were registered at a general practice at the start of follow up.

The treated population were those eligible for treatment with sotrovimab, paxlovid or molnupiravir, as defined by the NHS Commissioning Policy eligibility criteria for Covid-19 antivirals and nMAB in the community (11, 12), and applied by the centre dispensing treatment. This required individuals to i) have a SARS-CoV-2 infection confirmed by a positive polymerase chain reaction (PCR) or lateral flow test, ii) be symptomatic with Covid-19, and iii) belong to at least one of the following high risk cohorts: Down’s syndrome, solid cancer, haematological disease or stem cell transplant, renal disease, liver disease, immune-mediated inflammatory disorders, primary immune deficiencies, HIV/AIDS, solid organ transplant, or rare neurological conditions, and iv) have received an antiviral or nMAB during the study period. Although having symptomatic Covid-19 was part of the NHS Commissioning Policy eligibility criteria, we could not confirm this requirement in our analysis due to difficulties in determining symptom status using the data. Similarly, there were some treated individuals who we could not identify as belonging to a high-risk cohort using EHR data in the OpenSAFELY platform.

We identified an eligible but untreated population, defined as those with Covid-19 who were entitled to treatment (as set out in i to iii of the inclusion criteria for the treated patients above) but who never received antivirals or nMAB. For the untreated population, high-risk cohort allocation was defined by NHS Digital logic and associated code lists in EHR data in the OpenSAFELY platform (10)., allowing multiple categories for each patient. To ensure that the untreated population were not hospitalised when testing SARS-CoV-2 positive we excluded patients admitted to hospital before or on the date of their positive test. We also excluded individuals discharged on or 1 day prior to their positive SARS-CoV-2 test to ensure the positive test did not reflect a continued Covid-19 spell from a hospital admission. Detailed definitions and code lists are available online (https://www.opencodelists.org).

### Exposure

The exposure of interest was treatment with sotrovimab, paxlovid or molnupiravir in the Covid-19 therapeutics dataset within 5 days of a positive Covid-19 test. Patients were excluded if they had treatment records for any other nMAB or antiviral agents for Covid-19 before or on the same date as receiving sotrovimab, paxlovid or molnupiravir.

We emulated a hypothetical target trial. Our analyses used a per-protocol approach, where individuals in the treatment population were considered at risk from the start date of their treatment record. Individuals in the untreated population were considered at risk from 2-days after the date of their positive Covid-19 test. This 2-day delay was calculated as the median delay between a positive Covid-19 test and treatment initiation in the treatment group. As this was a safety analysis, the pre-protocol approach ensured that individuals in the treatment group could only contribute exposure time after treatment had been initiated. This approach provides a more reliable estimate of the true effect of treatment, and these exposure windows were chosen to minimise the risk of immortal time bias. An intention to treat analysis, whereby all individuals were considered at risk from the date of their positive Covid-19 test, was performed in sensitivity analyses. In all analyses, individuals were considered at risk until either: the outcome event date; 28 days after the start of treatment; the start of a second nMAB or antiviral agent in the treatment group; patient deregistration; or study end date (25^th^ July 2023); whichever occurred first.

### Outcomes

The outcomes of interest were pre-specified AEs, grouped into i) Summary of Product Characteristics (SmPC) AEs ii) drug reaction AEs and iii) immune-mediated AEs. SmPC AEs were events reported during the drug trial programme or listed in the drug’s SmPCs. The European Medicines Agency (EMA) publishes a SmPC for each drug on the European market, based on submissions from the drug manufacturer with patient-level data, alongside data gathered from spontaneous reporting by regulators or the marketing authorisation holder. For sotrovimab these included: diverticulitis, rash, contact dermatitis, bronchospasm, hypersensitivity, infusion-related reactions, and anaphylaxis (13); for paxlovid: dysgeusia, diarrhoea and nausea and vomiting (14), and for molnupiravir: dizziness, headache, diarrhoea, nausea and vomiting, rash and urticaria (15). Drug reaction AEs included anaphylaxis, angioedema, steven-Johnson syndrome, toxic epidermal necrolysis, hypersensitivity, and infusion-related reaction. Although hypersensitivity, infusion-related reactions and anaphylaxis are listed in the SmPCs for sotrovimab, these were grouped under drug reaction AEs. Immune-mediated AEs were examined after observational data suggested a safety signal with sotrovimab (16). Immune mediated AEs included rheumatoid arthritis, psoriatic arthritis, axial spondylarthritis, systemic lupus erythematosus and inflammatory bowel disease. These were identified if it was the first ever code for the condition, to ensure capture of a new event and not a flare of an underlying condition. For example, if a patient had a code for ‘rheumatoid arthritis’ during the at-risk period, this was only captured as an adverse event it the patient had no prior code for “rheumatoid arthritis” in their records. If they had a prior code for rheumatoid arthritis, it was assumed that the most recent coded event may refer to a flare of their underlying condition.

All AEs were extracted from three data sources: primary care record (SNOMED CT and CTV3 codes), Accident and Emergency (SNOMED CT codes) and hospital admission (ICD-10 codes) within Secondary Use Services (SUS) data. A hospitalisation code was only included if it was the primary cause of admission. Details of definitions and code lists are included in the supplementary 6. Events were defined as serious if the event resulted in hospitalisation.

### Covariates

Potential confounding factors were selected a priori and extracted at baseline. These included: age, sex, ethnic group, NHS region of general practice, index of multiple deprivation (IMD, grouped into five categories, derived from postcodes to reflect socioeconomic status), body mass index, covid-19 vaccination status (unvaccinated, and one, two, or three or more vaccinations), 10 high risk cohort categories (allowing multiple categories for each patient), and other comorbidities (diabetes, hypertension, chronic cardiac disease and chronic respiratory disease). Individuals with missing information for ethnic group, IMD, and BMI were included as an unknown category for each variable.

### Statistical analysis

Baseline characteristics of individuals by treatment status were tabulated and tested for statistically significant imbalance using chi squared, Mann–Whitney or t-tests, as appropriate. For each of the four patient groups (treated with paxlovid, sotrovimab or molnupiravir and untreated), 29-day event rates per 1000 patient-years with 95% confidence intervals (95% CIs) were calculated for individual AEs. To minimise the risk of disclosive data, frequencies of baseline characteristics, AE numbers and person exposure time were rounded to midpoint 6 (17). This method ensures that values between 1-6 are mapped to ‘3’. This ‘3’ is not a true 3 and is only a label for all numbers ranging between 1 and 6. The same process is applied to values between 7-12, which are mapped to ‘9’, and so on. Compared to other rounding options, rounding to the midpoint 6 improves preserves non-zero counts, improves precision and reduces bias.

An analysis of AEs in a pre-pandemic period was performed for a historical comparison, using the same cohort of individuals from the main analysis. This observation time started four years prior to each individual’s exposure start time in the main analysis. A 29-day event rate was calculated.

A Cox proportional hazards model was used to compare the risk across the four patient groups (treated with paxlovid, sotrovimab or molnupiravir and untreated) and reported as hazard ratios (HRs). The untreated population was chosen as the referent for comparison. In the Cox model, events were grouped into i) SmPC, ii) drug and iii) immune-mediated AEs. All SmPC AEs were examined, acknowledging that certain AEs were specific for one drug and not listed in another drug’s SmPC. Assumptions for each Cox model were tested using Nelson-Aalen plots and scaled Schoenfeld residuals.

### Sensitivity analyses

A propensity score (PS) model was used as an alternative method to account for confounding bias. The PS weighting balances the distributions of relevant covariates between the untreated population and those receiving either sotrovimab, paxlovid or molnupiravir. This was estimated as the probability of being treated by binary logistic regression of the actual treatment allocation using baseline covariates. Three logistic regression models were fitted, one for each drug. Baseline covariates included: age, sex, ethnic group, index of multiple deprivation (five categories), NHS region of general practice, vaccination status, body mass index, diabetes, hypertension, chronic cardiac and respiratory diseases. An inverse probability of treatment weights was estimated for patients in each drug group compared to the untreated group (supplementary 1).

As part of our per protocol approach, we conducted a sensitivity analysis where individuals in the untreated population were considered at risk from their positive Covid-19 test without incorporating a 2-day delay.

An intention to treat approach was also examined, whereby all individuals were considered at risk from the date of their positive Covid-19 test. In addition to this sensitivity analysis, a time-varying Cox model was employed, where individuals in the treatment group could initially contribute person-time to the untreated arm between their Covid-19 positive test and treatment initiation, and then contribute to the treatment group after treatment initiation.

Lastly, we performed two further sensitivity analyses, the first restricting the cohort to those who did not have a contraindication to paxlovid treatment, and the second, restricting the treated cohort to those who could be identified as eligible using NHS Digital logic and associated code lists in the OpenSAFELY platform (as used for identifying eligibility in the untreated population).

### Software and reproducibility

Data management was performed with Python, with analysis carried out with Stata 17. Code for data management and analysis, as well as codelists, are archived online (https://github.com/opensafely/Safety-Sotrovimab-Paxlovid-Molnupiravir).

### Patient and public involvement

Neither patients nor the public were involved in developing the research question and study, or in the design, management, or interpretation of this study.

## Results

### Patient characteristics

Between 16^th^ December 2021 and 26^th^ June 2023, there were 147,099 high-risk individuals with Covid-19 in the community who met the study inclusion criteria; of whom were in the exposure arms; sotrovimab (n=14,865), paxlovid (n=13,779) and molnupiravir (n=8,805), and 109,653 in the but untreated population. The most frequently represented high risk cohorts were immune-mediated inflammatory disorders on immunosuppression (37,641, 26%), and solid organ cancers (33,681, 23%) (table 1).

**Table 1.** Baseline characteristics of patients with Covid-19 treated with sotrovimab, paxlovid, molnupiravir and in the untreated population.

|  | Untreated<br>(n=109,653) |  | Sotrovimab group<br>(n=14865) |  | Paxlovid group<br>(n=13779) |  | Molnupiravir group<br>(n=8805) |  | Total<br>(n=147099) |  |
| --- | --- | --- | --- | --- | --- | --- | --- | --- | --- | --- |
| Age (years, mean (SD))* | 61 | 49-74 | 58 | 46-69 | 56 | 45-67 | 61 | 49-73 | 61 | 48-73 |
| Women | 59481 | 54.2 | 8697 | 58.5 | 8931 | 64.8 | 5025 | 57.1 | 82137 | 55.8 |
| Ethnicity |  |  |  |  |  |  |  |  |  |  |
| White | 97809 | 90.6 | 13719 | 92.9 | 12879 | 94.6 | 8205 | 94.1 | 132615 | 91.4 |
| Black | 3315 | 3.1 | 201 | 1.4 | 165 | 1.2 | 141 | 1.6 | 3819 | 2.6 |
| South Asian | 4359 | 4 | 549 | 3.7 | 357 | 2.6 | 225 | 2.6 | 5481 | 3.8 |
| Mixed | 1299 | 1.2 | 141 | 1 | 105 | 0.8 | 87 | 1 | 1641 | 1.1 |
| Other | 1167 | 1.1 | 159 | 1.1 | 111 | 0.8 | 63 | 0.7 | 1497 | 1 |
| Most deprived | 18207 | 17 | 1779 | 12.3 | 1341 | 10 | 903 | 10.5 | 22233 | 15.5 |
| Region (NHS)*: |  |  |  |  |  |  |  |  |  |  |
| East | 25317 | 23.1 | 4263 | 28.8 | 3435 | 25 | 3267 | 37.2 | 36285 | 24.7 |
| East Midlands | 18507 | 16.9 | 3531 | 23.8 | 3273 | 23.9 | 795 | 9 | 26109 | 17.8 |
| London | 5055 | 4.6 | 741 | 5 | 555 | 4 | 411 | 4.7 | 6765 | 4.6 |
| Northeast | 5763 | 5.3 | 801 | 5.4 | 267 | 1.9 | 105 | 1.2 | 6939 | 4.7 |
| Northwest | 9705 | 8.9 | 1197 | 8.1 | 1479 | 10.8 | 525 | 6 | 12909 | 8.8 |
| Southeast | 7749 | 7.1 | 723 | 4.9 | 1173 | 8.6 | 981 | 11.2 | 10629 | 7.2 |
| South West | 17619 | 16.1 | 2127 | 14.3 | 1737 | 12.7 | 1551 | 17.6 | 23037 | 15.7 |
| West Midlands | 3825 | 3.5 | 663 | 4.5 | 171 | 1.2 | 141 | 1.6 | 4797 | 3.3 |
| Yorkshire | 15873 | 14.5 | 777 | 5.2 | 1623 | 11.8 | 1017 | 11.6 | 19293 | 13.1 |
| High risk cohorts: |  |  |  |  |  |  |  |  |  |  |
| Down's syndrome* | 2433 | 2.2 | 195 | 1.3 | 309 | 2.2 | 207 | 2.4 | 3147 | 2.1 |
| Solid cancer | 28095 | 25.6 | 2631 | 17.7 | 1323 | 9.6 | 1635 | 18.6 | 33681 | 22.9 |
| Haematological disease | 9759 | 8.9 | 2529 | 17 | 1011 | 7.3 | 1281 | 14.6 | 14589 | 9.9 |
| Liver disease | 9765 | 8.9 | 849 | 5.7 | 153 | 1.1 | 519 | 5.9 | 11289 | 7.7 |
| Renal disease* | 10113 | 9.2 | 2727 | 18.3 | 39 | 0.3 | 1311 | 14.9 | 14187 | 9.6 |
| IMID | 26949 | 24.6 | 5109 | 34.4 | 2247 | 16.3 | 3333 | 37.9 | 37641 | 25.6 |
| Immunosuppression* | 20037 | 18.3 | 2787 | 18.8 | 2307 | 16.7 | 1725 | 19.6 | 26853 | 18.3 |
| HIV/AIDS* | 333 | 0.3 | 267 | 1.8 | 27 | 0.2 | 237 | 2.7 | 861 | 0.6 |
| Solid organ transplant* | 7599 | 6.9 | 2271 | 15.3 | 201 | 1.5 | 939 | 10.7 | 11007 | 7.5 |
| Rare neurological disease | 14505 | 13.2 | 1827 | 12.3 | 2217 | 16.1 | 1065 | 12.1 | 19617 | 13.3 |
| Chronic respiratory disease | 21351 | 19.5 | 3339 | 22.5 | 2313 | 16.8 | 2055 | 23.3 | 29061 | 19.8 |
| Chronic cardiac disease | 20217 | 18.4 | 2613 | 17.6 | 1065 | 7.7 | 1719 | 19.5 | 25611 | 17.4 |
| Diabetes | 26535 | 24.2 | 3531 | 23.8 | 2091 | 15.2 | 2205 | 25.1 | 34365 | 23.4 |
| Hypertension | 48315 | 44.1 | 6993 | 47.1 | 4071 | 29.5 | 4347 | 49.4 | 63729 | 43.3 |
| BMI |  |  |  |  |  |  |  |  |  |  |
| Underweight | 2823 | 2.7 | 231 | 1.6 | 219 | 1.7 | 165 | 2 | 3435 | 2.4 |
| Normal | 33741 | 32.2 | 4251 | 29.8 | 4401 | 33.2 | 2451 | 29.1 | 44847 | 31.9 |
| Overweight | 35337 | 33.7 | 4659 | 32.7 | 4317 | 32.6 | 2883 | 34.2 | 47193 | 33.6 |
| Obese | 32829 | 31.3 | 5115 | 35.9 | 4305 | 32.5 | 2937 | 34.8 | 45183 | 32.1 |
| Dementia | 3507 | 3.2 | 99 | 0.7 | 69 | 0.5 | 153 | 1.7 | 3831 | 2.6 |
| Serious mental illness | 1719 | 1.6 | 171 | 1.2 | 87 | 0.6 | 123 | 1.4 | 2103 | 1.4 |
| Care home resident | 4923 | 4.5 | 69 | 0.5 | 117 | 0.8 | 225 | 2.6 | 5343 | 3.6 |
| Vaccination status: |  |  |  |  |  |  |  |  |  |  |
| None | 4533 | 4.1 | 189 | 1.3 | 195 | 1.4 | 153 | 1.7 | 5073 | 3.4 |
| One | 1905 | 1.7 | 141 | 0.9 | 153 | 1.1 | 81 | 0.9 | 2283 | 1.6 |
| Two | 9075 | 8.3 | 609 | 4.1 | 393 | 2.9 | 405 | 4.6 | 10479 | 7.1 |
| Three | 60357 | 55 | 7407 | 49.8 | 4923 | 35.7 | 3981 | 45.2 | 76671 | 52.1 |
| Four or more | 33783 | 30.8 | 6519 | 43.9 | 8115 | 58.9 | 4179 | 47.5 | 52587 | 35.8 |
| Prior Covid-19 | 9555 | 8.7 | 999 | 6.7 | 1119 | 8.1 | 549 | 6.2 | 12231 | 8.3 |

Data are number (%) unless indicated otherwise. SD=standard deviation. To reduce the risk of disclosure, all counts have been rounded to midpoint 6 and percentages have been calculated using these rounded values; as such column totals might differ from the sum of the individual variables. Ethnic group, index of multiple deprivation and body mass index had 2043, 3807 and 6429 missing values, respectively.

There were differences between the treatment groups (table 1), with a younger cohort in the sotrovimab and paxlovid groups compared with the molnupiravir and untreated populations [median age 58 years (IQR 46 to 69) with sotrovimab, 56 (45-67) with paxlovid, 61 (48-73) with molnupiravir and 61 (49-74) in the untreated populations]. The untreated population had greater levels of deprivation [17% in the most deprived IMD quintile versus 10% to 12% among the treatment arms], greater proportion with prior Covid-19 infection [8.7% versus 6.2% to 8.1% among the treatment arms], lower vaccine uptake [4% unvaccinated versus 1.3% to 1.7% in the treatment arms and 31% with 4 or more vaccinations versus 43%-59% among the treatment arms] and more likely to be a care home resident [5% versus 1-3% among the treatment arms]. Compared with the other treatment arms, the paxlovid group had less cardiac disease [8% compared with 18-21%], respiratory disease [17% compared with 20-23%] and diabetes [15% compared with 24-25%], and the molnupiravir group were more likely to be a care home resident [2.6% compared with 0.5-0.8%] .

### Adverse events as listed in the drug’s SmPC

There were 1413 SmPC AEs, of which 345 resulted in hospitalisation. The 29-day rates of SmPC AEs were highest with sotrovimab compared with the other groups [sotrovimab 0.44 per 1000 patient-years (95% CI 0.38-0.51), paxlovid 0.39 (95% CI 0.33-0.45), molnupiravir 0.35 (95% CI 0.28-0.43) and the untreated population 0.36 (95% CI 0.33-0.38)] (figure 1). The rates of serious SmPC AEs, defined as those resulting in hospitalisation, were similar across the groups (supplementary 2). The most frequent SmPC AEs were nausea and vomiting, diarrhoea, and headache. Among the treated population, rash was more frequent in the sotrovimab group [0.09 (95% CI 0.07-0.12)] compared with paxlovid [0.05 (95% CI 0.03-0.08)] and molnupiravir [0.04 (95% CI 0.02-0.07)] whilst headache was more frequent in the paxlovid group [0.11 (95% CI 0.08-0.15)] compared with sotrovimab [0.08 (95% CI 0.05-0.11)] and molnupiravir [0.08 (95% CI 0.05-0.13)].

**Figure 1.**
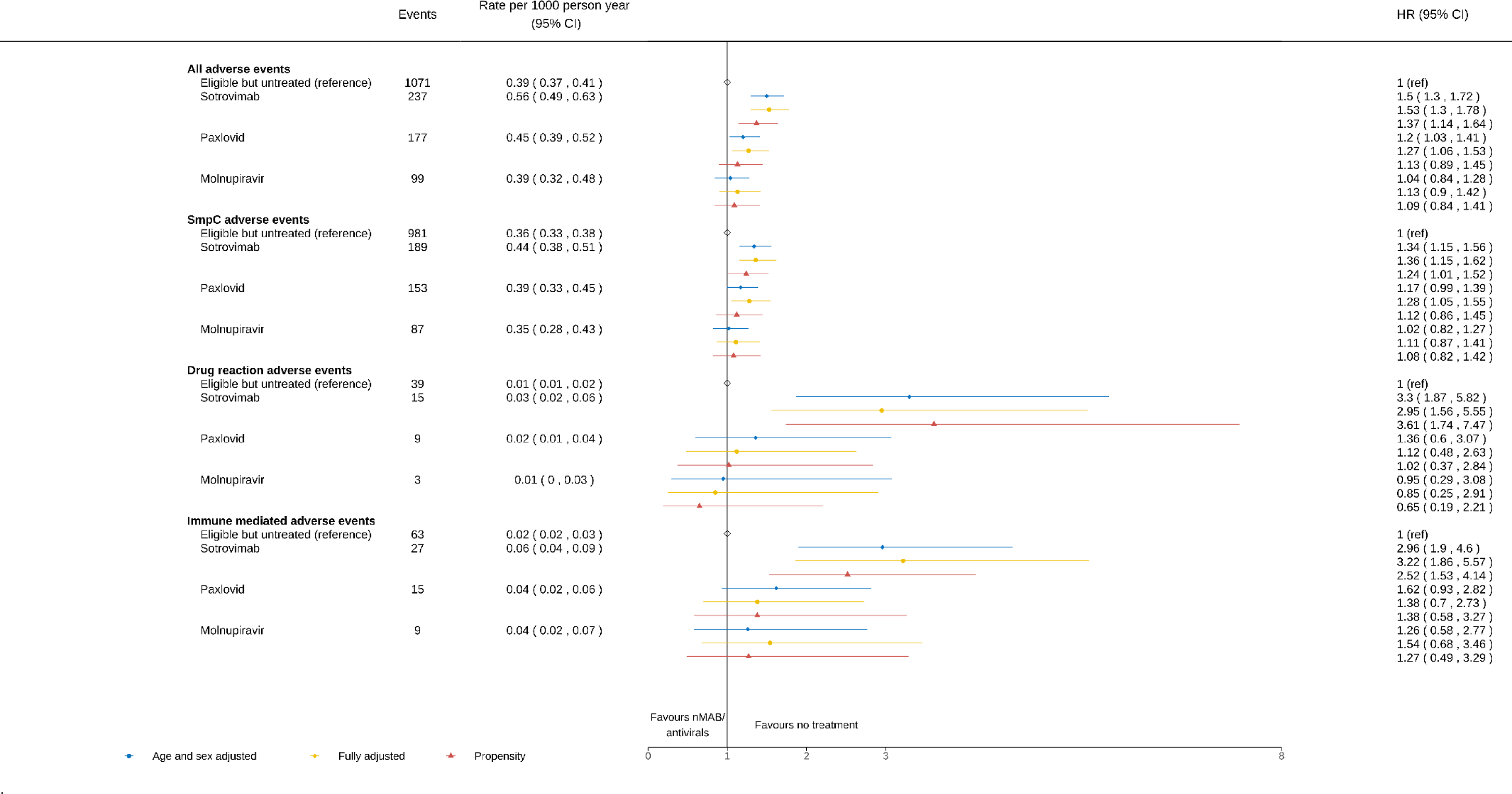
Comparing the risk of adverse events (SmPC, drug related and immune-mediated) during the 28 days of follow-up between patients treated with sotrovimab, paxlovid or molnupiravir, compared with the untreated population. Legend: Hazard ratio (95% confidence interval) for adverse events (SmPC, drug and immune mediated). *Fully adjusted model;* further adjusted for ethnic group, index of multiple deprivation (five categories), Sustainability Transformation Partnerships code of their registered GP surgery, vaccination status, body mass index, diabetes, hypertension, chronic cardiac and respiratory diseases, and the 10 high risk cohort categories. *Propensity model;* adjusted for age, sex, ethnic group, index of multiple deprivation (five categories), Sustainability Transformation Partnerships code of their registered GP surgery, vaccination status, body mass index, diabetes, hypertension, chronic cardiac and respiratory diseases. Propensity score model used the same baseline covariates as the adjusted model, but did not including the 10 high risk cohort categories. To reduce the risk of disclosure, all counts have been rounded to midpoint 6 and rates have been calculated using these rounded values.

In the analyses of pre-pandemic records performed for comparative purposes, there were similar numbers of SmPC AEs to that recorded in the primary analysis (1431 compared to 1413). The 29-day rate of SmPC AEs were also comparable; 0.34 (95% CI 0.32-0.36) in the pre-pandemic analysis and 0.37 (95% CI 0.35-0.39) in the primary analysis. The number of SmPC AEs leading to hospitalisation were also similar. There were however differences in the frequency of individual SmPC events between the two periods; with only 3 dysgeusia events compared with 45 in the primary analysis and 159 nausea and vomiting events compared with 369 in the primary analysis. Conversely, headache and rash were more frequent in the pre-pandemic analysis (table 2).

**Table 2.** Rates of all adverse events (AEs) during the study period and the historical comparative pre-pandemic period.

|  | All Study Population |  |  | Comparative Pre-Pandemic Records |  |  |
| --- | --- | --- | --- | --- | --- | --- |
|  | Events | Person Exp | Rate (95% CI)* | Events | Person Exp | Rate (95% CI)* |
| **SmPC AEs** | 1413 | 3824145 | 0.37 (0.35, 0.39) | 1431 | 4244373 | 0.34 (0.32, 0.36) |
| Diverticulitis | 99 | 3849507 | 0.03 (0.02, 0.03) | 99 | 4264503 | 0.02 (0.02, 0.03) |
| Diarrhoea | 351 | 3844881 | 0.9 (0.08, 0.10) | 309 | 4261137 | 0.07 (0.06, 0.08) |
| Taste | 45 | 3849609 | 0.01 (0.01, 0.02) | 3 | 4265865 | 0.00 (0.00, 0.00) |
| Rash | 201 | 3848403 | 0.05 (0.05, 0.06) | 303 | 4261143 | 0.07 (0.06, 0.08) |
| Bronchospasm | 0 | 3851139 | 0.00 (0.00, 0.00) | 3 | 4265841 | 0.00 (0.00, 0.00) |
| Contact dermatitis | 9 | 3851037 | 0.00 (0.00, 0.00) | 21 | 4265547 | 0.00 (0.00, 0.01) |
| Dizziness | 141 | 3848367 | 0.04 (0.03, 0.04) | 189 | 4263039 | 0.04 (0.04, 0.05) |
| Nausea and vomiting | 369 | 3843927 | 0.10 (0.09, 0.11) | 159 | 4263519 | 0.04 (0.03, 0.04) |
| Headache | 279 | 3844821 | 0.07 (0.06, 0.08) | 345 | 4260915 | 0.08 (0.07, 0.09) |
| **Drug reaction AEs** | 63 | 3850089 | 0.02 (0.01, 0.02) | 75 | 4264683 | 0.02 (0.01, 0.02) |
| Anaphylaxis | 9 | 3850971 | 0.00 (0.00, 0.01) | 9 | 4265739 | 0.00 (0.00, 0.00) |
| Drug reactions | 57 | 3850227 | 0.01 (0.01, 0.02) | 63 | 4264815 | 0.01 (0.01, 0.02) |
| **Immune mediated AEs** | 117 | 3849591 | 0.03 (0.03, 0.04) | 267 | 4261731 | 0.06 (0.06, 0.07) |
| Rheumatoid arthritis | 33 | 3850761 | 0.01 (0.01, 0.01) | 81 | 4264557 | 0.02 (0.02, 0.02) |
| SLE | 3 | 3851115 | 0.00 (0.00, 0.00) | 9 | 4265721 | 0.00 (0.00, 0.00) |
| Psoriasis | 21 | 3850905 | 0.01 (0.00, 0.01) | 39 | 4265307 | 0.01 (0.01, 0.01) |
| Psoriatic Arthritis | 9 | 3851043 | 0.00 (0.00, 0.00) | 33 | 4265367 | 0.01 (0.01, 0.01) |
| AxSpa | 21 | 3850887 | 0.01 (0.00, 0.01) | 27 | 4265439 | 0.01 (0.00, 0.01) |
| IBD | 27 | 3850701 | 0.01 (0.00, 0.01) | 87 | 4264611 | 0.02 (0.02, 0.03) |

Rate is 29-day event rates per 1000 patient-years with 95% confidence intervals. SLE: Systemic Lupus Erythematosus; AxSpA: Axial spondylarthritis; IBD Inflammatory bowel disease. To reduce the risk of disclosure, all counts and person exposure have been rounded to midpoint 6 and rates have been calculated using these rounded values

In the Cox proportional hazards model, compared with the untreated population, there was an increased risk of SmPC AEs with sotrovimab [adjusted HR 1.36 (95% CI 1.15-1.62)] and paxlovid [adjHR 1.28 (95% CI 1.05-1.55)]. This was not seen with molnupiravir [adjHR 1.11 (95% CI 0.87-1.41)] or when restricting to SmPC AEs that resulted in hospitalisation (supplementary 3). The propensity model demonstrated similar findings, although the increased risk of SmPC AEs with paxlovid was present but no longer statistically significant [propensity adjHR 1.12 (95% CI 0.86-1.45)] (figure 1).

### Drug reaction adverse events

There were 63 drug reaction AEs, including 9 cases of anaphylaxis of which 3 resulted in an inpatient hospitalisation. The 29-day rate of drug reaction was highest in the sotrovimab arm and lowest in the molnupiravir and untreated population [sotrovimab 0.03 (95% CI 0.02-0.06), paxlovid 0.02 (95% CI 0.01-0.04), molnupiravir 0.01 (95% CI 0.00-0.03) and the untreated population 0.01 (95% CI 0.01-0.02)] (figure 1). There were numerically more drug reaction AEs in the pre-pandemic analysis compared with the primary analysis (75 versus 63) although the 29-day rates were similar between the two periods (table 2).

In the Cox proportional hazards model, compared with the untreated population, there was an increased risk of drug reaction AEs with sotrovimab [adjHR 2.95 (95% CI 1.56-5.55)]. This was not seen with paxlovid [adjHR 1.12 (95% CI 0.48-2.63)] or molnupiravir [adjHR 0.85 (95% CI 0.25-2.91)]. The propensity model demonstrated similar findings to the main analysis, although the risk with sotrovimab was greater [propensity adjHR 3.61 (95% CI 1.74-7.47)] (figure 1).

### Immune-mediated adverse events

There were 117 immune-mediated AEs; rheumatoid arthritis and inflammatory bowel disease were the most frequent events (33 and 27 events, respectively). There were 51 immune mediated AEs requiring hospitalisation. The 29-day rate of all immune-mediated AEs was highest in the sotrovimab arm and lowest in the untreated population [sotrovimab 0.06 (95% CI 0.04-0.09), paxlovid 0.04 (95% CI 0.02-0.06), molnupiravir 0.04 (95% CI 0.02-0.07) and the untreated population 0.02 (95% CI 0.02-0.03)] (figure 1).

There were 267 immune-mediated AEs in the pre-pandemic period. The number of rheumatoid arthritis, inflammatory bowel disease, psoriasis, psoriatic arthritis, and SLE events were 2 to 3-fold higher than that recorded in the primary analysis. The 29-day rate of immune-mediated AEs were higher in the pre-pandemic period: 0.06 (95% CI 0.06-0.07)] compared with 0.03 (95% CI 0.03-0.04) in the primary analysis, whilst the rate of immune-mediated AEs leading to hospitalisation was 0.02 (95% CI 0.02-0.03) compared with 0.01 (95% CI 0.01-0.02) in the primary analysis (table 2 & supplementary 4).

In the Cox proportional hazards model, compared with the untreated population, there was an increased risk of immune-mediated AEs with sotrovimab [adjHR 3.22 (95% CI 1.86-5.57)]. This was not seen with paxlovid [adjHR 1.38 (95% CI 0.70-2.73)] or molnupiravir [adjHR 1.54 (95% CI 0.68-3.46)]. When restricting to immune-mediated AEs that resulted in hospitalisation, there was no increased risk with treatment (supplementary 3). The propensity model demonstrated similar findings to the main analysis, although the risk with sotrovimab was smaller [propensity adjHR 2.52 (95% CI 1.53-4.14)] (figure 1).

### Sensitivity analysis

In the per protocol approach, without incorporating a 2-day delay after positive Covid-19 test in the untreated group, the risk of SmPC AEs with sotrovimab and paxlovid was no longer statistically significant [sotrovimab adj HR 1.11 (95% CI 0.94-1.30) and paxlovid adjHR 1.03 (95% CI 1.85-1.25)]. The risk of drug reactions and immune-mediated AEs with sotrovimab were still present [adjHR 2.19 (95% CI 1.17-4.12)] and [adjHR 2.78 (95% CI 1.63-4.73)] respectively (figure 2).

**Figure 2.**
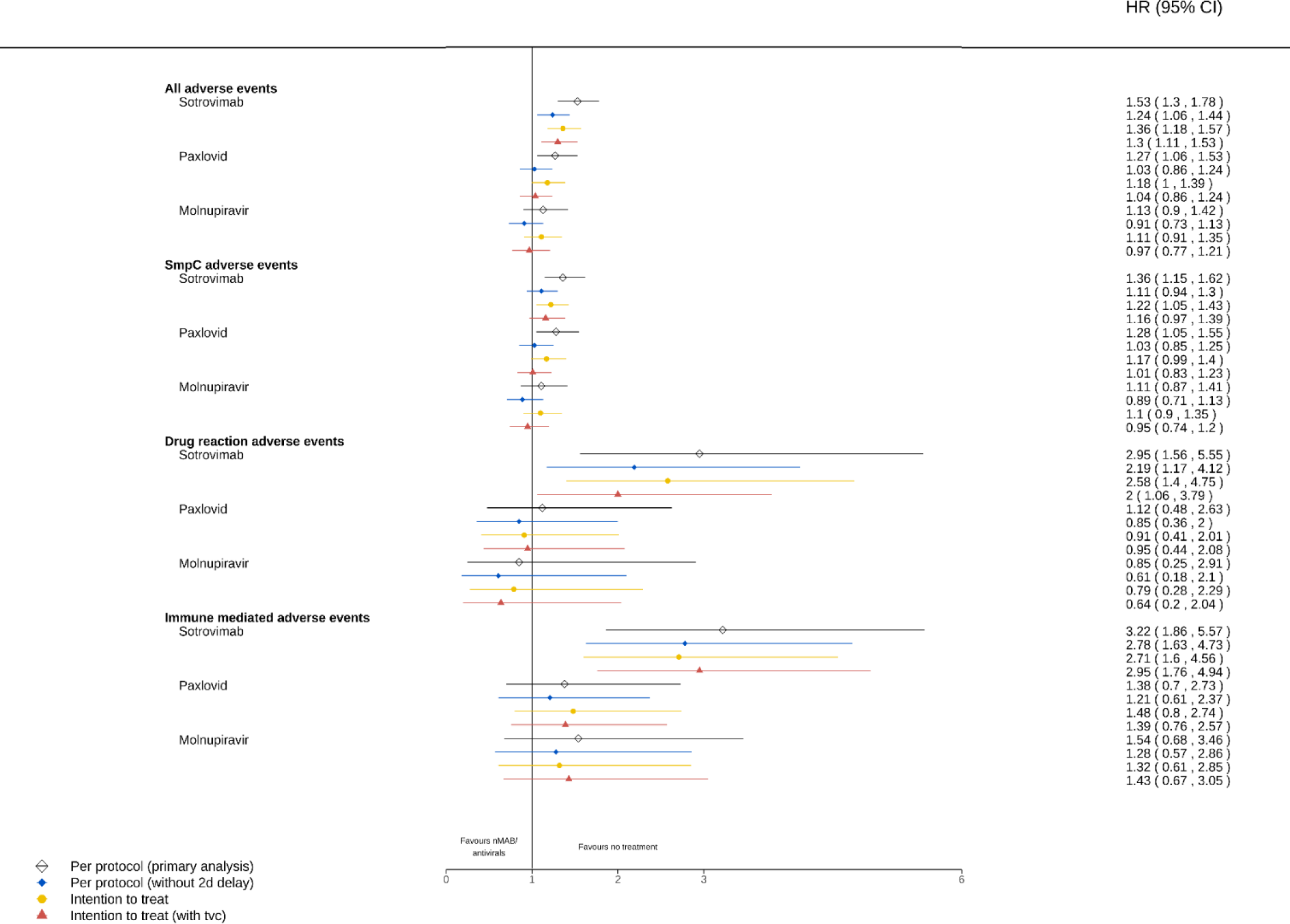
Sensitivity analyses comparing the risk of adverse events (SmPC, drug-related and immune-mediated) between patients treated with sotrovimab paxlovid and molnupiravir, compared with the untreated population, using different exposure windows and a time-varying Cox model. Legend: Hazard ratio (95% confidence interval) for adverse events (SmPC, drug and immune mediated). *Per protocol approach* (primary analysis fully adjusted model): Individuals in the treatment population were considered at risk from the start date of their treatment record. Individuals in the untreated population were considered at risk from 2-days after the date of their positive Covid-19 test. *Per protocol approach (without 2-day delay):* Individuals in the untreated population were considered at risk from the date of their positive Covid-19 test without incorporating 2 day delay. *Intention to treat approach:* All individuals considered at risk from the date of their positive Covid-19 test. *Intention to treat approach (with a time varying Cox model):* All individuals considered at risk from the date of their positive Covid-19 test, but using a time-varying Cox model individuals in the treatment group could initially contribute person-time to the untreated arm between their Covid-19 positive test and treatment initiation, and then contribute to the treatment group after treatment initiation.

In the intention-to-treat approach, whereby all individuals were considered at risk from the date of their positive Covid-19 test, the 29-day rates of AEs were similar to per protocol approach. The risks of SmPC, drug reaction and immune-mediated AEs with sotrovimab were still present, whilst the risk of SmPC AEs with paxlovid was no longer statistically significant (figure 2) (supplementary 5). In the time-varying Cox model, where individuals in the treatment group could initially contribute person-time to the untreated arm between their Covid-19 positive test and treatment initiation, and then contribute to the treatment group after treatment initiation, the risks of drug reaction and immune-mediated AEs with sotrovimab was still present, although the risk of SmPC AEs was no longer statistically significant (figure 2).

When the cohort was restricted to individuals without a contraindication to paxlovid (eGFR <30, liver disease, organ transplant and potential drug interactions) results were similar to those seen in the primary analysis (supplementary 6).

When the treated cohort was restricted to individuals who could be identified as eligible using NHS Digital logic and associated code lists alone, the risk of SmPC and drug reaction AEs with sotrovimab were no longer statistically significant [adj HR 1.02 (95% CI 0.84-1.25) and 1.69 (95% CI 0.78-3.66)]; however, the risk of immune-mediated AEs with sotrovimab was still present [adjHR 3.53 (95% CI 2.01-6.19)] (supplementary 7).

## Discussion

This national cohort study has examined pharmacovigilance of antivirals and nMABs in the prehospital treatment of Covid-19, using real-world data from primary and secondary care electronic health records via OpenSAFELY. In a population of over 160,000 high-risk individuals with Covid-19, 25% were prescribed treatment with either sotrovimab, paxlovid or molnupiravir by CMDUs across England. The untreated cohort comprised individuals who were identified as eligible but did not receive therapy. Overall, the rates of pre-specified AEs were low. Sotrovimab was associated with an increased risk of SmPC, drug-related and immune-mediated AEs, although rates of these events were similar or lower to that seen during a comparative historical period. These findings, particularly the risk of drug and immune-mediated AEs, remained robust in propensity score weighting analysis and in sensitivity analyses utilising a range of potential exposure start dates in both the treated and untreated populations.

Our current understanding of the safety of antivirals and nMABs has been derived from clinical trial data, which is often underpowered to detect rare AEs. Meta-analyses allow the quantitative synthesis of safety data across multiple trials, and these have been largely reassuring and in keeping with the findings from our study. A meta-analysis of molnupiravir RCTs did not find an increase in the risk of serious and non-serious AEs compared with placebo (18). A meta-analysis of 13 paxlovid studies (including 4 RCTs) did report an increased risk of overall AEs (OR 2.31, 95% CI 1.36–3.94) compared with placebo and standard of care, however these findings were no longer seen in subgroup analyses limiting to clinical trial data (19). In contrast to our results, an analysis of 5 RCTs examining nMABs, of which only one trial assessed sotrovimab, reported a reduction in serious AEs (OR: 0.37 95% CI: 0.19– 0.72), although this may be explained by most SAEs being hospitalizations for Covid-19–related causes. No increase in the risk of non-serious AEs or in several predetermined SmPC events was reported (20). It is important to consider the limitations of meta-analyses when interpreting safety signals, including heterogeneity of included studies with variability in safety reporting, incomplete or missing trial data, publication bias and limited generalisability of the trial cohort to a real-world population.

There are numerous real-world studies analysing the effectiveness of sotrovimab, paxlovid and molnupiravir. These analyses rarely focus on drug safety. There are few safety analyses, and these are limited by small sample sizes, reducing the ability to detect rare events (21-24). A real-world safety analysis across all of England, but limited to hospital admission data, found no association between sotrovimab and 26 prespecified adverse outcomes (16). When splitting the 28-day risk period into narrower windows, an increased risk of hospitalisation with rheumatoid arthritis and SLE were described in the early post exposure period. This is difficult to interpret for two reasons: firstly it is not clear if these were primary or secondary causes for admission, the latter may incorrectly identify a comorbid condition rather than a true cause for hospitalisation; and, secondly, these admissions were not clearly defined as new diagnoses, and may have represented an individual with established disease presenting with a flare, which for both rheumatoid arthritis and SLE can be triggered by infection. In our study, we report an increased risk of immune-mediated AEs with sotrovimab. These events were identified using both primary and secondary care data. AEs were defined as new diagnoses and, if resulting in hospitalisation, the event was the primary cause for admission, indicating with greater confidence that an immune safety signal exists. This should however be interpreted alongside the much lower rates of immune-mediated AEs compared with that seen during the historical pre-pandemic period. This is supported by the decrease in the incidence of immune-mediated conditions reported during the pandemic, which coincided with rising rates of Covid-19 (25).

A key strength of our study is the scale, level of detail and completeness of primary care EHR data with linkage to inpatient hospital records within the OpenSAFELY-TPP platform. Identifying an eligible but untreated control population has provided a basis for a pragmatic safety comparator and allowed us to try to account for selection bias and confounding. The duration of the study and the use of near real-time EHR data permitted the analysis of all three therapies, despite national guidance on the choice of drug evolving over time with the emergence of evidence on therapeutic efficacy.

There are also several limitations of our study. Firstly, one of the challenges of pharmacovigilance using EHR data is how to define relevant AEs. For this study, we compiled definitions based on AEs listed in the drugs’ SmPC, in addition to outcomes of interest identified during the trial programme or from observational analyses. However, pre-specifying events of interest is reliant upon there being adequate time after the licensing of new therapies for rare events to become evident, as well as ensuring a thorough review of existing evidence before defining AEs. It remains possible that there are safety signals that remain undetected due to these limitations. Additionally, the detection of AEs relies upon coding of clinical events by clinicians or clinical coding teams and may be subject to misclassification bias. ICD-10 codes in secondary care are less granular than SNOMED codes in primary care, which could also contribute to misclassification. Furthermore, some clinical events in general practice are recorded in free text, rather than as a coded diagnosis; this information is not captured in OpenSAFELY, due to the potential for re-identification. One must also consider the time lag between clinical presentation, diagnosis, and primary care coding. This is particularly relevant for immune-mediated AEs, where the initial onset of symptoms may have predated the drug exposure. This would affect drug groups equally and could have little effect on comparisons between treated and untreated populations. However, as immune-mediated AEs are rare, a small increase in events could have a major impact. Conversely, in secondary care, we only included AEs that were the primary cause for admission, as coding for secondary admission diagnoses is often less robust. As all individuals had Covid-19, it is possible that Covid-19 was defined as the primary admission diagnosis, despite the AE being the driver for hospitalisation.

Secondly, the identification of belonging to a high-risk cohort was different for the untreated and treated populations. For the untreated population, high-risk cohort allocation was defined by code lists in EHR data, which is sensitive but not specific in identifying high-risk individuals (10). For the treated population, high-risk cohort allocation was defined by the centres dispensing treatment, and it is not possible to examine this eligibility using EHR data.

Thirdly, in order to emulate a trial design, we employed both per protocol and intention to treat approaches. For our main analysis we used the per protocol approach, where the exposure windows for our treated and untreated populations commenced at different times (i.e., at treatment start date in the treated population, and at Covid-19 test date plus a 2-day delay in the untreated population). This was chosen to help mitigate the risk of immortal time bias. However, this approach is also imperfect, as we use future information (being treated after a Covid-19 positive test) to stratify our population into treated and untreated groups, potentially introducing bias. Importantly, our sensitivity analyses utilising an intention to treat approach with Covid-19 test date as the exposure start date for all populations yielded similar findings to our primary analysis, particularly in regards to the risk of drug and immune-mediated AEs with sotrovimab. Furthermore, sensitivity analyses using the per protocol approach without a delay in the exposure start date in the untreated population demonstrated similar findings. Using these different exposure windows increases our confidence in these estimates, particularly in respect to drug and immune-mediated AE risk with sotrovimab. Alternative approaches to account for immortal time bias., such as sequential trial emulation or cloning are also available but we did not explore these as we felt they would not provide substantial benefits, e.g., for a sequential trial design most individuals are eligible at only a single interval so little is gained in terms of power and precision.

Fourthly, the analysis of pre-pandemic records provided important context for the observed event rates and aided in the interpretation of safety signals. However, it is plausible that in this historic period, immune-mediated AEs were disproportionately high, as analyses were being performed in the high-risk cohort, of which immune-mediated conditions is one of the risk groups. As such, the analysis of pre-pandemic records may have captured their initial immune-mediated diagnosis.

Finally, there is the possibility of residual confounding, related to the differences between the treatment arms. For example, the untreated cohort may not have presented to CMDUs to start antiviral or nMAB treatment, despite being at high risk of Covid-19 complications. There are likely unmeasured factors that contributed to whether these individuals sought treatment, which in turn may have influenced subsequent clinical outcomes. This bias could have favoured the risk of AEs in the untreated group, who may have been less likely to present to a health worker with an AE. In contrast, it is possible that some individuals in the untreated cohort did present to CMDUs but were not prescribed treatment as their Covid infection was mild or asymptomatic, or their comorbidities were not considered severe enough to meet eligibility. This bias could also have favoured the risk of AEs in the untreated group, who may have been a ‘healthier’ cohort. There are additional differences within the treated group, with unmeasured factors influencing a clinicians’ choice of therapeutic agent and contributing to confounding by indication. It is difficult to appreciate exactly how this bias has influenced our results.

The timeliness, granularity, and scale of the data available via OpenSAFELY offers unprecedented opportunity to carry out rapid observational work to inform the roll-out of new treatments such as monoclonal antibodies and antiviral therapies. Compared with RCT and industry-led post-marketing trials, EHRs provide a larger and more diverse patient population with near-real time identification of emerging safety signals, permitting a comprehensive understanding of medication safety in real-world healthcare settings. There is an exciting opportunity to integrate real-world pharmacovigilance practice directly into healthcare data analysis. OpenSAFELY is now being used across more than 20 organisations and we encourage applications from more including regulators engaged in this area such as the Medicines and Healthcare products Regulatory Agency (MHRA) We believe this will bring substantial benefits with the continuous and comprehensive surveillance of medication effects across vast and diverse patient populations.

In conclusion, this study has shown an acceptable safety profile with sotrovimab, paxlovid and molnupiravir. Sotrovimab is associated with an increased risk of AEs, although event rates were lower than that seen in a comparative historical period. This work serves as a proof of principle study demonstrating the use of EHR data in the examination of pharmacovigilance.

## Data Availability

If requested, use the following: Access to the underlying identifiable and potentially re-identifiable pseudonymised electronic health record data is tightly governed by various legislative and regulatory frameworks, and restricted by best practice. The data in the NHS England OpenSAFELY COVID-19 service is drawn from General Practice data across England where TPP is the data processor.
TPP developers initiate an automated process to create pseudonymised records in the core OpenSAFELY database, which are copies of key structured data tables in the identifiable records. These pseudonymised records are linked onto key external data resources that have also been pseudonymised via SHA-512 one-way hashing of NHS numbers using a shared salt. University of Oxford, Bennett Institute for Applied Data Science developers and PIs, who hold contracts with NHS England, have access to the OpenSAFELY pseudonymised data tables to develop the OpenSAFELY tools.
These tools in turn enable researchers with OpenSAFELY data access agreements to write and execute code for data management and data analysis without direct access to the underlying raw pseudonymised patient data, and to review the outputs of this code. All code for the full data management pipeline from raw data to completed results for this analysis and for the OpenSAFELY platform as a whole is available for review at github.com/OpenSAFELY.
The data management and analysis code for this paper was led by Katie Bechman and contributed to by James Galloway.

## Acknowledgements

We are very grateful for all the support received from the TPP Technical Operations team throughout this work, and for generous assistance from the information governance and database teams at NHS England / NHSX. This study uses electronic health records, data provided by patients and collected by the National Health Service as part of their care and support.

## Information governance and ethical approval

NHS England is the data controller of the NHS England OpenSAFELY COVID-19 Service; TPP is the data processor; all study authors using OpenSAFELY have the approval of NHS England^1^. This implementation of OpenSAFELY is hosted within the TPP environment which is accredited to the ISO 27001 information security standard and is NHS IG Toolkit compliant;^2^. Patient data has been pseudonymised for analysis and linkage using industry standard cryptographic hashing techniques; all pseudonymised datasets transmitted for linkage onto OpenSAFELY are encrypted; access to the NHS England OpenSAFELY COVID-19 service is via a virtual private network (VPN) connection; the researchers hold contracts with NHS England and only access the platform to initiate database queries and statistical models; all database activity is logged; only aggregate statistical outputs leave the platform environment following best practice for anonymisation of results such as statistical disclosure control for low cell counts^3^.

The service adheres to the obligations of the UK General Data Protection Regulation (UK GDPR) and the Data Protection Act 2018. The service previously operated under notices initially issued in February 2020 by the Secretary of State under Regulation 3(4) of the Health Service (Control of Patient Information) Regulations 2002 (COPI Regulations), which required organisations to process confidential patient information for COVID-19 purposes; this set aside the requirement for patient consent^4^. As of 1 July 2023, the Secretary of State has requested that NHS England continue to operate the Service under the COVID-19 Directions 2020^5^. In some cases of data sharing, the common law duty of confidence is met using, for example, patient consent or support from the Health Research Authority Confidentiality Advisory Group^6^. Taken together, these provide the legal bases to link patient datasets using the service. GP practices, which provide access to the primary care data, are required to share relevant health information to support the public health response to the pandemic and have been informed of how the service operates.

This study was approved by the Health Research Authority (REC Reference 20/LO/0651) and by the LSHTM Ethics Board (Reference 21863).

## Data access and verification

If requested, use the following: Access to the underlying identifiable and potentially re-identifiable pseudonymised electronic health record data is tightly governed by various legislative and regulatory frameworks, and restricted by best practice. The data in the NHS England OpenSAFELY COVID-19 service is drawn from General Practice data across England where TPP is the data processor.

TPP developers initiate an automated process to create pseudonymised records in the core OpenSAFELY database, which are copies of key structured data tables in the identifiable records. These pseudonymised records are linked onto key external data resources that have also been pseudonymised via SHA-512 one-way hashing of NHS numbers using a shared salt. University of Oxford, Bennett Institute for Applied Data Science developers and PIs, who hold contracts with NHS England, have access to the OpenSAFELY pseudonymised data tables to develop the OpenSAFELY tools.

These tools in turn enable researchers with OpenSAFELY data access agreements to write and execute code for data management and data analysis without direct access to the underlying raw pseudonymised patient data, and to review the outputs of this code. All code for the full data management pipeline — from raw data to completed results for this analysis — and for the OpenSAFELY platform as a whole is available for review at github.com/OpenSAFELY.

The data management and analysis code for this paper was led by Katie Bechman and contributed to by James Galloway.

## Contributions

KB, JBG, BZ and SN conceptualised the study. KB, JBG, MDR, SN, BZ, AG and BMK developed the methodology. KB, JBG, AG and SN conducted the formal analysis. KB, BZ and MDR developed the diagnostic codelists. BG, BMK, AMe, RMS, SB and AG developed the software. KB wrote the original draft of the manuscript. All authors reviewed and edited the manuscript. All authors read and approved the final manuscript. KB and JBG are the guarantors for the Article, and accept full responsibility for the work and the conduct of the study. KB, JBG, and AG had full access to all the data in the study, and accessed and verified the data. All authors had final responsibility for the decision to submit for publication.

