## Supplementary for "The safety of antivirals and neutralising monoclonal antibodies used in prehospital treatment of Covid-19"

**Supplementary 1.** The propensity score (PS) was generated by using a logistic regression model, regressing treatment status (either sotrovimab and untreated, paxlovid and untreated or molnupiravir and untreated) on observed baseline characteristics including age, sex, ethnic group, index of multiple deprivation (five categories), NHS region of general practice, vaccination status, body mass index, diabetes, hypertension, chronic cardiac and respiratory diseases. The propensity score was then used to balance covariates using the inverse probability of treatment weighting (IPTW) method. The balancing of the cohorts using the weighted model was examined by comparing standardised differences between cohorts.

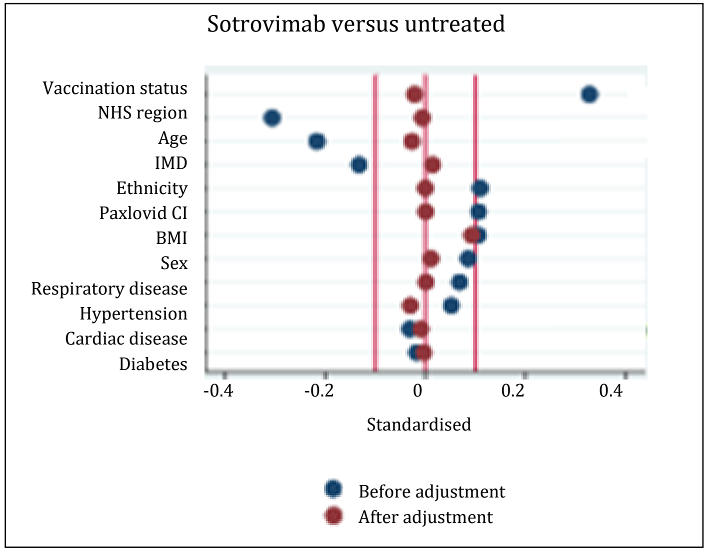


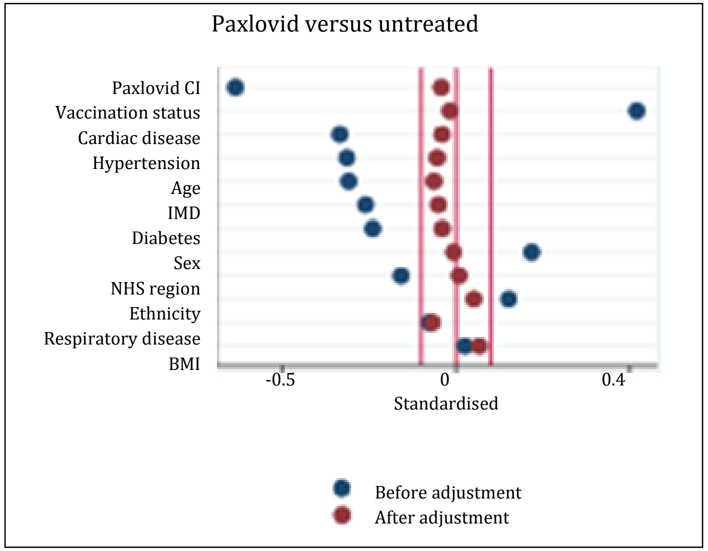


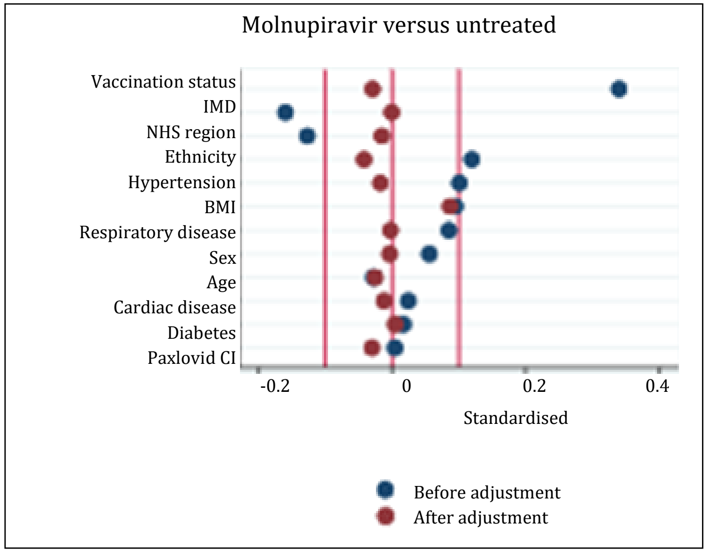

Supplementary 2. Rates of specific adverse events (AEs) by treatment group during the study period.

| | **Untreated** | | | **Sotrovimab group** | | | **Paxlovid group** | | | **Molnupiravir group** | | |
|---|---|---|---|---|---|---|---|---|---|---|---|---|
| | Event | PerExp | Rate (95% CI) * | Event | PerExp | Rate (95% CI) * | Event | PerExp | Rate (95% CI) * | Event | PerExp | Rate (95% CI) * |
| **All AEs** | 1071 | 2750607 | 0.39 (0.37, 0.41) | 237 | 425679 | 0.56 (0.49, 0.63) | 177 | 394227 | 0.45 (0.39, 0.52) | 99 | 251181 | 0.39 (0.32, 0.48) |
| **SmPC** | 981 | 2751951 | 0.36 (0.33, 0.38) | 189 | 426381 | 0.44 (0.38, 0.52) | 153 | 394491 | 0.39 (0.33, 0.45) | 87 | 251319 | 0.35 (0.28, 0.44) |
| Diverticulitis | 75 | 2770599 | 0.03 (0.02, 0.03) | 9 | 429231 | 0.02 (0.01, 0.04) | 9 | 397005 | 0.02 (0.01, 0.04) | 9 | 252669 | 0.04 (0.02, 0.07) |
| Diarrhoea | 243 | 2767203 | 0.09 (0.08, 0.10) | 39 | 428745 | 0.09 (0.07, 0.12) | 33 | 396543 | 0.08 (0.06, 0.12) | 27 | 252387 | 0.11 (0.07, 0.16) |
| Taste | 21 | 2770881 | 0.01 (0.00, 0.01) | 9 | 429141 | 0.02 (0.01, 0.04) | 9 | 396879 | 0.02 (0.01, 0.04) | 3 | 252705 | 0.01 (0.00, 0.03) |
| Rash | 135 | 2770077 | 0.05 (0.04, 0.06) | 39 | 428799 | 0.09 (0.07, 0.12) | 21 | 396885 | 0.05 (0.03, 0.08) | 9 | 252639 | 0.04 (0.02, 0.07) |
| Bronchospasm | 0 | 2771871 | 0.00 (0.00, 0.00) | 0 | 429345 | 0.01 (0.00, 0.01) | 0 | 397137 | 0.00 (0.00, 0.01) | 0 | 252789 | 0.00 (0.00, 0.01) |
| Contact dermatitis | 9 | 2771787 | 0.00 (0.00, 0.01) | 0 | 429345 | 0.00 (0.00, 0.01) | 0 | 397137 | 0.00 (0.00, 0.01) | 3 | 252771 | 0.01 (0.00, 0.03) |
| Dizziness | 93 | 2769813 | 0.03 (0.03, 0.04) | 21 | 429021 | 0.05 (0.03, 0.07) | 15 | 396855 | 0.04 (0.02, 0.06) | 9 | 252675 | 0.04 (0.02, 0.07) |
| Nausea & vomiting | 285 | 2766171 | 0.10 (0.09, 0.12) | 45 | 428607 | 0.10 (0.08, 0.14) | 27 | 396669 | 0.07 (0.05, 0.10) | 15 | 252483 | 0.06 (0.04, 0.10) |
| Headache | 177 | 2767275 | 0.06 (0.06, 0.07) | 33 | 428793 | 0.08 (0.05, 0.11) | 45 | 396357 | 0.11 (0.08, 0.15) | 21 | 252399 | 0.08 (0.05, 0.13) |
| **Drug reaction** | 39 | 2771313 | 0.01 (0.01, 0.02) | 15 | 428991 | 0.03 (0.02, 0.06) | 9 | 397053 | 0.02 (0.01, 0.04) | 3 | 252729 | 0.01 (0.00, 0.03) |
| Anaphylaxis | 3 | 2771799 | 0.00 (0.00, 0.00) | 3 | 429273 | 0.01 (0.00, 0.02) | 0 | 397137 | 0.00 (0.00, 0.02) | 3 | 252759 | 0.01 (0.00, 0.03) |
| Drug reaction | 33 | 2771385 | 0.01 (0.01, 0.02) | 15 | 429033 | 0.03 (0.02, 0.06) | 9 | 397053 | 0.02 (0.01, 0.04) | 3 | 252753 | 0.01 (0.00, 0.03) |
| **Immune mediated** | 63 | 2771001 | 0.02 (0.02, 0.03) | 27 | 428937 | 0.06 (0.04, 0.09) | 15 | 396945 | 0.04 (0.02, 0.06) | 9 | 252705 | 0.04 (0.02, 0.07) |
| Rheumatoid | 21 | 2771673 | 0.01 (0.00, 0.01) | 3 | 429261 | 0.01 (0.00, 0.02) | 3 | 397083 | 0.01 (0.00, 0.02) | 3 | 252747 | 0.01 (0.00, 0.03) |
| SLE | 3 | 2771871 | 0.00 (0.00, 0.00) | 3 | 429321 | 0.01 (0.00, 0.02) | 0 | 397137 | 0.00 (0.00, 0.02) | 0 | 252789 | 0.00 (0.00, 0.01) |

| | Untreated | | | Sotrovimab group | | | Paxlovid group | | | Molnupiravir group | | |
|---|---|---|---|---|---|---|---|---|---|---|---|---|
| | Event | PerExp | Rate (95% CI) * | Event | PerExp | Rate (95% CI) * | Event | PerExp | Rate (95% CI) * | Event | PerExp | Rate (95% CI) * |
| Psoriasis | 15 | 2771733 | 0.01 (0.00, 0.01) | 3 | 429267 | 0.01 (0.00, 0.02) | 3 | 397113 | 0.01 (0.00, 0.02) | 0 | 252789 | 0.00 (0.00, 0.01) |
| Psoriatic Arthritis | 9 | 2771823 | 0.00 (0.00, 0.01) | 3 | 429303 | 0.01 (0.00, 0.02) | 0 | 397137 | 0.00 (0.00, 0.01) | 3 | 252783 | 0.01 (0.00, 0.03) |
| AxSpA | 9 | 2771757 | 0.00 (0.00, 0.01) | 3 | 429249 | 0.01 (0.00, 0.02) | 3 | 397089 | 0.01 (0.00, 0.02) | 3 | 252783 | 0.01 (0.00, 0.03) |
| IBD | 9 | 2771631 | 0.00 (0.00, 0.01) | 9 | 429249 | 0.01 (0.00, 0.02) | 9 | 397059 | 0.02 (0.01, 0.04) | 3 | 252759 | 0.01 (0.00, 0.03) |

PerExp: Person exposure.
*Rate is 29-day event rates per 1000 patient-years with 95% confidence intervals.
To reduce the risk of disclosure, all counts and person exposure have been rounded to midpoint 6 and rates have been calculated using these rounded values.

SLE: Systemic Lupus Erythematosus; AxSpA: Axial spondylarthritis; IBD Inflammatory bowel disease.

**Supplementary 3.** Comparing risk of <u>serious</u> adverse events (SmPC and immune mediated) during the 28 days of follow-up between patients treated with sotrovimab paxlovid and molnupiravir compared to the untreated population.

| | Events | Rate per 1000 person year (95% CI) |
|---|---|---|
| **All serious adverse events** | | |
| Eligible but untreated (reference) | 267 | 0.1 ( 0.09 , 0.11 ) |
| Sotrovimab | 57 | 0.13 ( 0.1 , 0.17 ) |
| Paxlovid | 45 | 0.11 ( 0.08 , 0.15 ) |
| Molnupiravir | 27 | 0.11 ( 0.07 , 0.16 ) |
| **Serious SmpC adverse events** | | |
| Eligible but untreated (reference) | 237 | 0.09 ( 0.08 , 0.1 ) |
| Sotrovimab | 45 | 0.1 ( 0.08 , 0.14 ) |
| Paxlovid | 39 | 0.1 ( 0.07 , 0.13 ) |
| Molnupiravir | 21 | 0.08 ( 0.05 , 0.13 ) |
| **Serious Immune mediated adverse events** | | |
| Eligible but untreated (reference) | 27 | 0.01 ( 0.01 , 0.01 ) |
| Sotrovimab | 9 | 0.02 ( 0.01 , 0.04 ) |
| Paxlovid | 9 | 0.02 ( 0.01 , 0.04 ) |
| Molnupiravir | 3 | 0.01 ( 0 , 0.03 ) |

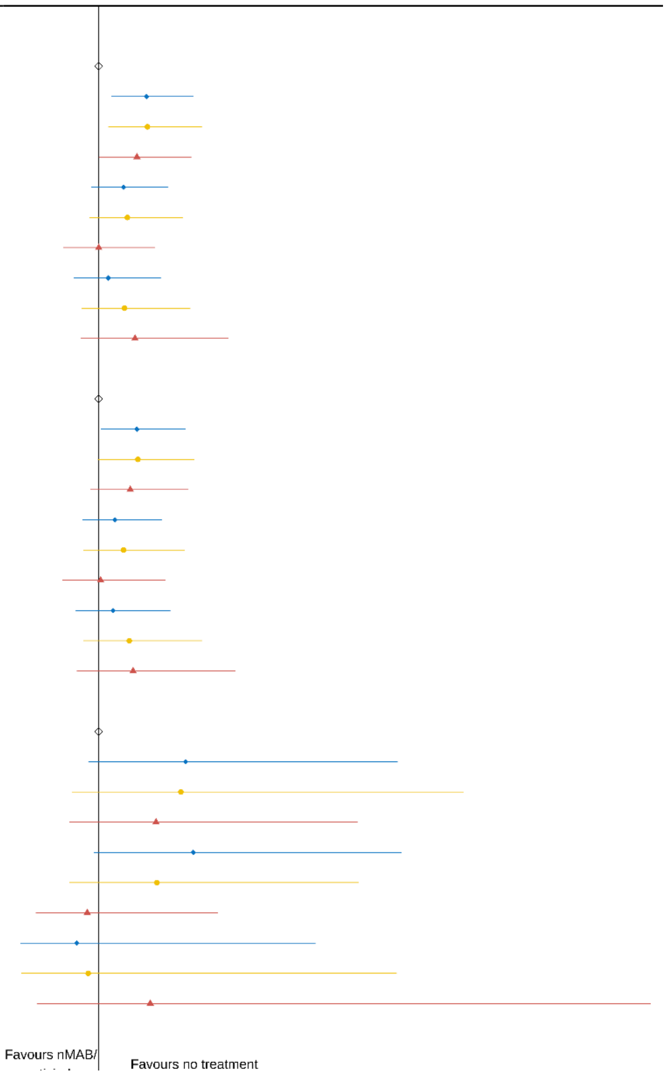


Hazard ratio (95% confidence interval) for adverse events (SmPC and immune mediated). Serious drug reactions not presented as very low events number. To reduce the risk of disclosure, all counts have been rounded to midpoint 6 and rates have been calculated using these rounded values.

Age and sex adjusted model.

Fully adjusted model; further adjusted for ethnic group, index of multiple deprivation (five categories), Sustainability Transformation Partnerships code of their registered GP surgery, vaccination status, body mass index, diabetes, hypertension, chronic cardiac and respiratory diseases, and the 10 high risk cohort categories. Propensity model; adjusted for age, sex, ethnic group, index of multiple deprivation (five categories), Sustainability Transformation Partnerships code of their registered GP surgery, vaccination status, body mass index, diabetes, hypertension, chronic cardiac and respiratory diseases.

Propensity score model used a baseline covariate including age, sex, ethnic group, index of multiple deprivation (five categories), NHS region of general practice, vaccination status, body mass index, diabetes, hypertension, chronic cardiac and respiratory diseases.S

Supplementary 4. Rates of adverse events (AEs) during the study period and the historical comparative pre-pandemic period.

*Rate is 29-day event rates per 1000 patient-years with 95% confidence intervals. SLE: Systemic Lupus Erythematosus; AxSpA: Axial spondylarthritis; IBD Inflammatory bowel disease

| | All Study Population | | | Comparative Pre-Pandemic Records | | |
|---|---|---|---|---|---|---|
| | Events | Person Exp | Rate (95% CI) * | Events | Person Exp | Rate (95% CI) * |
| **Serious SmPC AEs** | 345 | 3845643 | 0.09 (0.08, 0.10) | 321 | 4260981 | 0.08 (0.07, 0.08) |
| Diverticulitis | 63 | 3850263 | 0.02 (0.01, 0.02) | 69 | 4264887 | 0.02 (0.01, 0.02) |
| Diarrhoea | 123 | 3849195 | 0.03 (0.03, 0.04) | 135 | 4263747 | 0.03 (0.03, 0.04) |
| Taste | 0 | 3851169 | 0.00 (0.00, 0.00) | 0 | 4265871 | 0.00 (0.00, 0.00) |
| Rash | 33 | 3850851 | 0.01 (0.01, 0.01) | 21 | 4265541 | 0.00 (0.00, 0.01) |
| Contact dermatitis | 3 | 3851145 | 0.00 (0.00, 0.00) | 21 | 4265547 | 0.00 (0.00, 0.01) |
| Dizziness | 9 | 3850923 | 0.00 (0.00, 0.00) | 15 | 4265655 | 0.00 (0.00, 0.01) |
| Nausea and vomiting | 93 | 3849399 | 0.02 (0.02, 0.03) | 21 | 4265517 | 0.00 (0.00, 0.01) |
| Headache | 21 | 3850869 | 0.01 (0.00, 0.01) | 45 | 4265283 | 0.01 (0.01, 0.01) |
| **Serious drug reaction AEs** | 3 | 3851067 | 0.00 (0.00, 0.00) | 3 | 4265829 | 0.00 (0.00, 0.00) |
| Anaphylaxis | 3 | 3851073 | 0.00 (0.00, 0.00) | 3 | 4265847 | 0.00 (0.00, 0.00) |
| Drug reactions | 3 | 3851157 | 0.00 (0.00, 0.00) | 3 | 4265853 | 0.00 (0.00, 0.0) |
| **Serious immune mediated AEs** | 51 | 3850617 | 0.01 (0.01, 0.02) | 99 | 4264395 | 0.00 (0.00, 0.01) |
| Rheumatoid arthritis | 15 | 3851025 | 0.00 (0.00, 0.01) | 21 | 4265517 | 0.00 (0.00, 0.00) |
| SLE | 3 | 3851157 | 0.00 (0.00, 0.00) | 3 | 4265829 | 0.00 (0.00, 0.00) |
| Psoriasis | 3 | 3851133 | 0.00 (0.00, 0.00) | 3 | 4265793 | 0.00 (0.00, 0.00) |
| Psoriatic Arthritis | 3 | 3851145 | 0.00 (0.00, 0.00) | 3 | 4265811 | 0.00 (0.00, 0.00) |
| AxSpa | 15 | 3850989 | 0.00 (0.00, 0.01) | 21 | 4265529 | 0.00 (0.00, 0.01) |
| IBD | 15 | 3851001 | 0.00 (0.00, 0.01) | 45 | 4265217 | 0.01 0.01, 0.01) |

To reduce the risk of disclosure, all counts and person exposure have been rounded to midpoint 6 and rates have been calculated using these rounded values.

Supplementary 5. Rates of all adverse events (AEs) calculated from per protocol approach (main analysis where treatment population were considered at risk from the start date of their treatment record and untreated population were considered at risk from 2-days after the date of their positive Covid-19 test) and intention to treat approach (sensitivity analysis where all individuals were considered at risk from the date of their positive Covid-19 test).

| | Per protocol (main analysis) | | | intention to treat approach (sensitivity analysis) | | |
|---|---|---|---|---|---|---|
| | Events | Person Exp | Rate (95% CI) * | Events | Person Exp | Rate (95% CI) * |
| **SmPC AEs** | 1413 | 3824145 | 0.37 (0.35, 0.39) | 1785 | 4093389 | 0.44 (0.42, 0.46) |
| Diverticulitis | 99 | 3849507 | 0.03 (0.02, 0.03) | 117 | 4121313 | 0.03 (0.02, 0.03) |
| Diarrhoea | 351 | 3844881 | 0.9 (0.08, 0.10) | 417 | 4116453 | 0.01 (0.09, 0.11) |
| Taste | 45 | 3849609 | 0.01 (0.01, 0.02) | 81 | 4121061 | 0.02 (0.02, 0.02) |
| Rash | 201 | 3848403 | 0.05 (0.05, 0.06) | 213 | 4120173 | 0.05 (0.05,0.06) |
| Bronchospasm | 0 | 3851139 | 0.00 (0.00, 0.00) | 3 | 4122939 | 0.00 (0.00, 0.00) |
| Contact dermatitis | 9 | 3851037 | 0.00 (0.00, 0.00) | 9 | 4122873 | 0.00 (0.00, 0.00) |
| Dizziness | 141 | 3848367 | 0.04 (0.03, 0.04) | 171 | 4120065 | 0.04 (0.04, 0.05) |
| Nausea and vomiting | 369 | 3843927 | 0.10 (0.09, 0.11) | 465 | 4115385 | 0.11 (0.10, 0.12) |
| Headache | 279 | 3844821 | 0.07 (0.06, 0.08) | 417 | 4115085 | 0.10 (0.09, 0.11) |
| **Drug reaction AEs** | 63 | 3850089 | 0.02 (0.01, 0.02) | 81 | 4121859 | 0.02 (0.02, 0.02) |
| Anaphylaxis | 9 | 3850971 | 0.00 (0.00, 0.01) | 9 | 4122807 | 0.00 (0.00, 0.00) |
| Drug reactions | 57 | 3850227 | 0.01 (0.01, 0.02) | 69 | 4121997 | 0.02 (0.01, 0.02) |
| **Immune mediated AEs** | 117 | 3849591 | 0.03 (0.03, 0.04) | 129 | 4121385 | 0.03 (0.03, 0.04) |
| Rheumatoid arthritis | 33 | 3850761 | 0.01 (0.01, 0.01) | 33 | 4122609 | 0.01 (0.01, 0.00) |
| SLE | 3 | 3851115 | 0.00 (0.00, 0.00) | 3 | 4122945 | 0.00 (0.00, 0.01) |
| Psoriasis | 21 | 3850905 | 0.01 (0.00, 0.01) | 27 | 4122723 | 0.01 (0.00, 0.01) |
| Psoriatic Arthritis | 9 | 3851043 | 0.00 (0.00, 0.00) | 15 | 4122849 | 0.00 (0.00, 0.01) |
| AxSpa | 21 | 3850887 | 0.01 (0.00, 0.01) | 21 | 4122705 | 0.01 (0.00, 0.01) |
| IBD | 27 | 3850701 | 0.01 (0.00, 0.01) | 39 | 4122519 | 0.01 (0.01, 0.01) |

*Rate is 29-day event rates per 1000 patient-years with 95% confidence intervals. SLE: Systemic Lupus Erythematosus; AxSpA: Axial spondylarthritis; IBD Inflammatory bowel disease. To reduce the risk of disclosure, all counts and person exposure have been rounded to midpoint 6 and rates have been calculated using these rounded values.

**Supplementary 6.** Sensitivity analyses: comparing risk of adverse events (SmPC, drug related and immune mediated) during the 28 days of follow-up between patients treated with sotrovimab paxlovid and molnupiravir compared to the untreated population, restricting to those without contraindication to paxlovid.

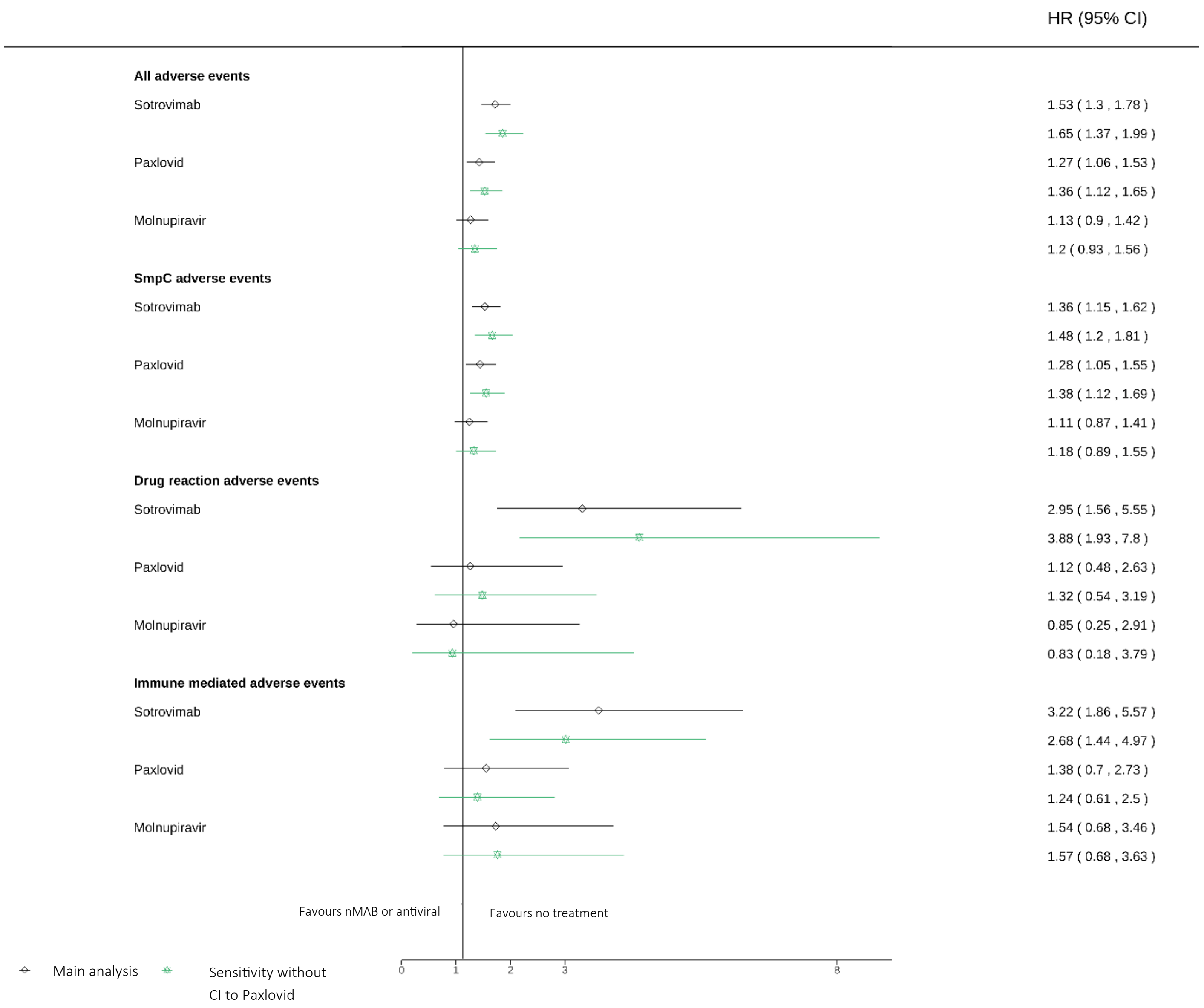

Supplementary 7: Sensitivity analyses: comparing risk of adverse events (SmPC, drug related and immune mediated) during the 28 days of follow-up between patients treated with sotrovimab paxlovid and molnupiravir compared to the untreated population, restricting the treated cohort to those who could be identified as eligible using NHS Digital logic and associated code lists in the OpenSAFELY platform (as used for identifying eligibility in the untreated population).

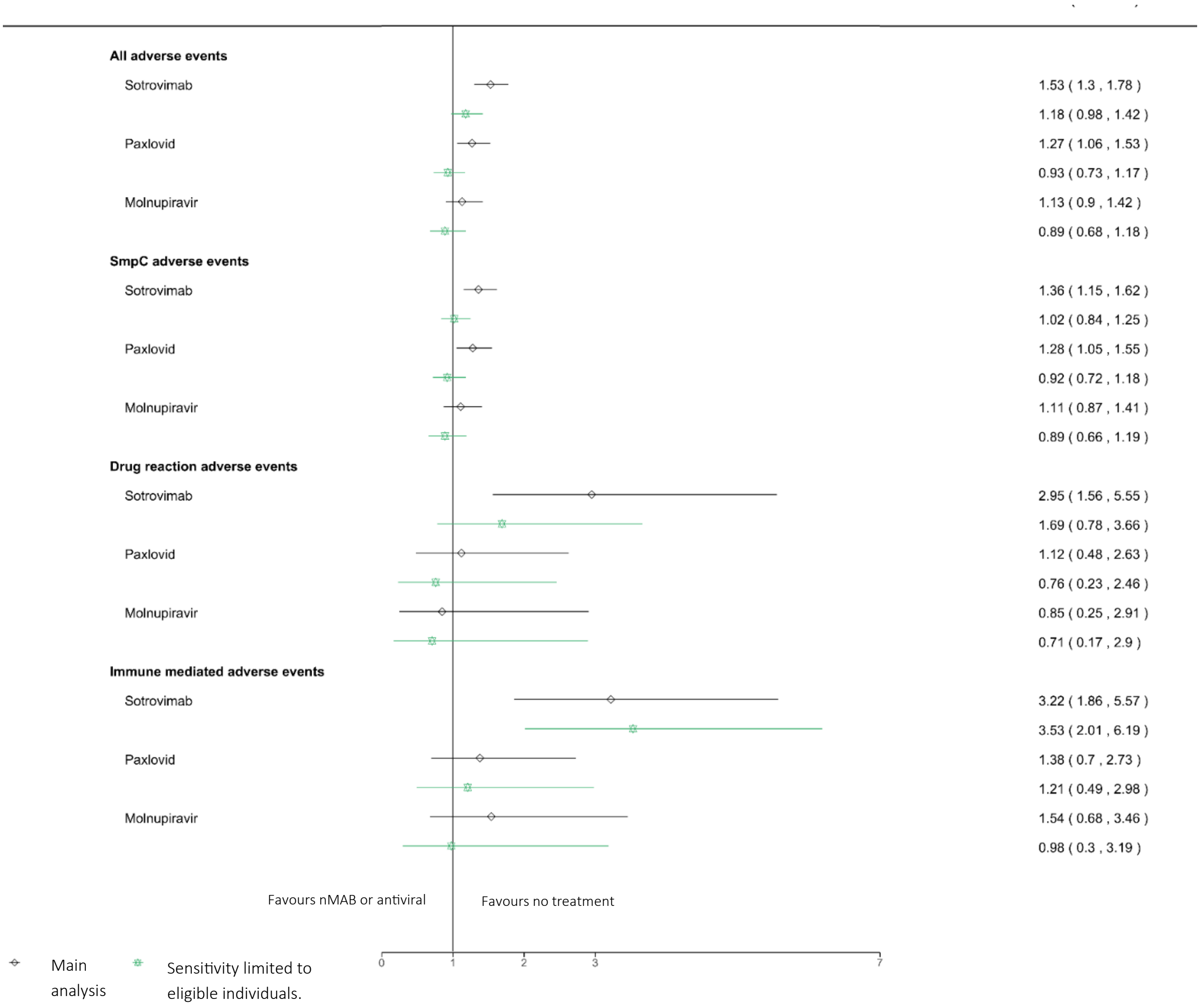
